# End-to-End Reliability of Automated Systems for Diagnostic Data Extraction: A Benchmark Study in Uro-Oncologic Evidence Synthesis

**DOI:** 10.64898/2025.12.24.25342959

**Authors:** Matthias May, Julio Ruben Rodas Garzaro, Anton Kravchuk, Christian Gilfrich, Gregor Donabauer, Samy Ateia, Udo Kruschwitz, Maximilian Haas, Christoph Eckl, Maximilian Burger

## Abstract

**Background:** Automated systems, including large language models, are increasingly used to support data extraction in diagnostic systematic reviews. However, their reliability, safety, and repeatability under realistic extraction conditions remain insufficiently characterized.

**Objective:** To benchmark the end-to-end reliability of automated systems for extracting diagnostic accuracy data from published uro-oncologic studies, with a focus on correctness, abstention behavior in non-derivable scenarios, repeatability across repeated runs, and operational efficiency.

**Methods:** This prospective, protocol-driven benchmarking study evaluates a purpose-built extraction system (MedNuggetizer) and three contemporary large language models. Systems are applied to a fixed corpus of published full-text PDFs and publicly available supplementary material reporting on Uromonitor and urine cytology for bladder cancer detection. A locked, uniform extraction prompt is used across all systems. The primary endpoint is dataset-run correctness, defined as either exact extraction of the complete 2-by-2 diagnostic table or correct declaration of non-derivability. A non-inferiority design with an exact one-sided binomial test is employed. Secondary endpoints include hallucination behavior on pre-specified sentinel datasets, repeatability across repeated runs, fidelity of derived diagnostic metrics, and execution time compared with human extraction.

**Results:** The study is powered for a non-inferiority margin of 5 percentage points relative to a predefined correctness threshold of 95 percent. Twenty independent runs per system are performed, yielding 320 dataset-run observations. Primary inference is conducted at the run level, with consensus-level results reported as supportive robustness analyses.

**Conclusions:** This protocol establishes a conservative and reproducible framework for evaluating automated systems used in diagnostic evidence synthesis. By integrating correctness, abstention, repeatability, and safety into a single end-to-end evaluation, the study addresses key methodological gaps in the clinical assessment of generative AI tools.

## 1. Background and rationale (revised)

### 1.1 Clinical and academic need

Diagnostic test evidence in uro-oncology is commonly synthesized through systematic reviews and diagnostic accuracy meta-analyses. The core unit of diagnostic accuracy is the 2×2 contingency table, defined by four integers: true positives (TP), false positives (FP), false negatives (FN), and true negatives (TN). From these values, investigators derive sensitivity, specificity, positive and negative predictive values, and overall accuracy. These metrics shape how diagnostic tests are perceived in routine care, how they enter surveillance pathways, and how they are discussed in multidisciplinary tumor boards. Data extraction is the highest-risk step in this chain. A single incorrect cell can materially distort downstream metrics, especially in modest cohorts, low-prevalence settings, or fragmented reporting. For a clinically active academic urology group, the practical question is not whether automated systems can assist, but whether they can be used without compromising evidentiary integrity.

*This protocol addresses a user-relevant, clinically anchored question:* Can contemporary large language models and a group-developed extraction system (MedNuggetizer) be used to extract diagnostic accuracy data from full-text PDFs and supplementary material in a manner that is reliable, reproducible, and safe for systematic review workflows?

### 1.2 What the methods literature shows, and what it does not

Recent work supports the premise that extraction performance is context dependent and that failure modes are systematic rather than random [1–5]. Key observations include the influence of reporting structure, the prevalence of omission and unsupported numeric assertions, and the fact that effective benchmark size is determined by the number of extraction decisions rather than the number of publications.

The literature also highlights a gap that is directly relevant to clinical evidence synthesis. Most evaluations focus on numeric accuracy when extraction is feasible, but provide less clarity on behavior in edge cases where the required 2×2 table is not derivable from the provided documents. For clinical workflows, correct abstention is not a limitation. It is a safety feature. Accordingly, a clinically meaningful benchmark should not only quantify correctness when extraction is possible, but also quantify safety behavior when extraction is not possible. This protocol operationalizes both components under locked prompting conditions.

### 1.3 Why MedNuggetizer requires clinical-grade validation

MedNuggetizer is a confidence-based information nugget extraction pipeline intended to support clinicians and researchers working with long-form medical documents [6]. Its design emphasizes repeated extraction and consolidation to reduce omission and stabilize outputs across runs. A clinical-grade validation must therefore address two questions that are not interchangeable:

1. End-to-end reliability in realistic document processing, including correct extraction when data are derivable and correct abstention when they are not.
2. Repeatability across repeated runs under the same locked prompting conditions, reflecting intraobserver variability in automated extraction.

This study evaluates four extraction systems, including MedNuggetizer as the primary system under validation and three Large Language Model (LLM)-based extraction pipelines as external comparators, using a locked diagnostic corpus. A human benchmark serves as the sole reference standard for document derivability and for correctness of extracted 2×2 cells. Systems are evaluated under identical prompting constraints, without external sources and without model-specific prompt optimization.

### 1.4 Why Uromonitor and urine cytology provide a suitable benchmark

The benchmark corpus is anchored in a PROSPERO-registered systematic review and diagnostic accuracy meta-analysis of the Uromonitor molecular urine assay, currently in peer review (*diagnostics-4082381*) [7]. The corpus comprises eight publications and supports extraction attempts for two index tests per publication: Uromonitor and urine cytology. This structure yields a set of datasets that resemble the real extraction workload of a diagnostic accuracy review, where the presence and derivability of a complete 2×2 table cannot be assumed in advance.

A key feature of this corpus is the presence of pre-specified non-derivable sentinel conditions. For the publication by Rubio-Briones et al., neither Uromonitor nor urine cytology 2×2 tables can be unambiguously derived from the PDF and publicly available supplementary material [8]. In addition, urine cytology is not available for Azawi et al. [9]. These cases allow an explicit safety evaluation because automated systems do not have access to unpublished author communications. In this setting, correct behavior is to declare non-derivability and avoid numeric output, rather than to generate plausible but unsupported integers.

### 1.5 Rationale for an integrated, clinically oriented evaluation

The intended publication is designed to be interpretable and actionable for urologists. Therefore, the protocol integrates three complementary layers into a single coherent evaluation:

1. End-to-end dataset-level correctness under locked prompting, reflecting the practical user question of whether a system can be trusted across heterogeneous reporting conditions.
2. Repeatability across 20 independent runs per system, quantifying intraobserver variability and the stability of extraction or abstention decisions.
3. Safety behavior on pre-specified sentinel non-derivable datasets, where any numeric emission is treated as unsafe.

*Together, these elements reflect the real decision faced by a urologic research group:* whether such systems can be integrated into evidence synthesis workflows without compromising validity, reproducibility, or safety.

## 2. Objectives, endpoints, hypotheses, and statistical analysis plan

### 2.1 Study objectives

#### Primary objective

To evaluate the end-to-end reliability of MedNuggetizer for diagnostic evidence synthesis when applied to full-text scientific articles and supplements under a locked, uniform prompt, reflecting the real-world user scenario in which derivability of diagnostic accuracy data is unknown a priori.

#### Secondary objectives

1. To evaluate whether contemporary large language models (LLMs) demonstrate non-inferior end-to-end reliability compared with the same benchmark under identical conditions.
2. To quantify safety behavior, specifically the absence of numeric hallucinations, on pre-specified non-derivable sentinel datasets.
3. To quantify within-system repeatability (intraobserver variability) across repeated runs for MedNuggetizer and each LLM.
4. To compare operational efficiency (execution time) of MedNuggetizer and LLMs versus human extraction.

### 2.2 Dataset structure and evaluation units

Eight peer-reviewed publications are evaluated. For each publication, two diagnostic tests are queried using locked prompts:

- Uromonitor
- Urine cytology

This yields 16 diagnostic datasets per run. Each system (MedNuggetizer and each evaluated LLM) is executed for R independent runs, each run initiated in a fresh session according to the protocol. Human benchmark extraction is performed once per dataset following the benchmark protocol.

#### Pre-specified sentinel non-derivable datasets

- Rubio-Briones et al.: Uromonitor and urine cytology (expected non-derivable) [8]
- Azawi et al.: urine cytology (expected non-derivable) [9]

These sentinel datasets are included in the primary end-to-end endpoint as correct abstention cases and are additionally evaluated in the safety analysis.

### 2.3 Primary endpoint: end-to-end dataset-run correctness

The primary endpoint is dataset-run correctness, defined for each diagnostic dataset in each run as follows:

- Benchmark-derivable datasets: Correct if and only if the system outputs a complete 2×2 contingency table with all four cells (TP, FP, FN, TN) exactly matching the human benchmark.
- Benchmark-non-derivable datasets: Correct if and only if the system outputs “Not derivable” and outputs no numeric values.

All 16 diagnostic datasets per run are included. The primary analysis therefore comprises N = 16 × R binary (correct/incorrect) evaluation units.

*This endpoint reflects the user-relevant question:* Does the system reliably extract correct diagnostic data when possible and reliably abstain when extraction is not possible?

### 2.4 Secondary endpoints

#### (1) Derivable-only numeric extraction performance

Restricted to benchmark-derivable datasets:

- Dataset-level exactness (all four cells correct)
- Cell-level exact-match accuracy for TP, FP, FN, and TN Reported descriptively with confidence intervals.

#### (2) Safety / hallucination endpoint

Restricted to sentinel non-derivable datasets:

- Unsafe behavior is defined as any numeric output when the correct behavior is “Not derivable”.
- Outcomes are summarized as hallucination rates with exact confidence intervals.

#### (3) Intraobserver variability / repeatability

Across R runs, repeatability is assessed on two levels:

- Output-category stability: agreement across runs for the three-category outcome (correct numeric extraction / correct abstention / incorrect or unsafe).
- Numeric stability (derivable datasets only): proportion of runs producing an identical complete 2×2 table.

#### (4) Operational efficiency

Per-run execution time (seconds), defined as the interval from run initiation to completion of protocol-conform output for all eight publications.

### 2.5 Hypotheses

Let *p* denote the probability of a correct dataset-run output under the primary endpoint definition.

#### H1 (primary; non-inferiority of MedNuggetizer)

MedNuggetizer is non-inferior to a reliability threshold of 95%, with a non-inferiority margin Δ = 5% points.

The null hypothesis is p ≤ 0.95, and the alternative hypothesis is p > 0.95.

#### H2 (secondary; non-inferiority of LLMs)

Each evaluated LLM is tested analogously against the same 95% threshold.

#### H3 (secondary; safety)

On sentinel non-derivable datasets, MedNuggetizer and the LLMs demonstrate correct abstention behavior with no numeric hallucinations.

#### H4 (secondary; repeatability)

Across repeated runs, MedNuggetizer demonstrates high within-system repeatability, exceeding that of single-pass LLM usage patterns.

#### H5 (secondary; operational efficiency)

Median execution time per run is shorter for MedNuggetizer and each LLM compared with human extraction.

### 2.6 Biometric planning and required number of runs

The study uses a fixed corpus of eight publications. Statistical power is therefore controlled by the number of repeated runs RRR.

#### Test parameters (primary endpoint)

- One-sided non-inferiority test
- α = 0.025
- The non-inferiority threshold was set at p₀ = 0.95
- Target assurance (power) ≥ 80%
- The conservative assumed true performance was set at p₁ = 0.98

#### Decision rule

A non-inferiority claim is made if the one-sided exact binomial probability under *p*_0_ = 0.95 of observing *X* or more correct dataset-runs is less than or equal to 0.025. With **R = 20 runs**:

- N = 16 × 20 = 320 dataset-run observations
- Critical value k = 312
- An observed accuracy of at least 97.5% is required
- Power is approximately 80% assuming p₁ = 0.98

Therefore, R = 20 runs is pre-specified to ensure a defensible non-inferiority design without reliance on optimistic assumptions.

### 2.7 Statistical analysis plan

#### Primary analysis (confirmatory)

- Outcome: dataset-run correctness (binary)
- Test: one-sided exact binomial test
- Significance level: α = 0.025
- Effect reporting: observed accuracy, exact one-sided 97.5% lower confidence bound

#### Secondary non-inferiority analyses (LLMs)

- Same binomial framework as H1
- Holm adjustment across LLM comparisons (family-wise α = 0.025)

#### Safety analysis

- Endpoint: hallucination (yes/no) on sentinel datasets
- Reporting: hallucination rate with exact 95% CI
- Between-system comparison: Fisher’s exact test (exploratory or Holm-adjusted)

#### Repeatability analysis

- Output-category agreement across runs: Fleiss’ κ with 95% CI per dataset
- Summary: median κ and IQR per system
- Between-system comparison: dataset-paired Wilcoxon signed-rank test
- Numeric stability (derivable datasets): Proportion of runs matching the modal 2×2 table Compared between systems using paired Wilcoxon signed-rank tests

#### Operational efficiency

- Per-run execution time
- Comparison to human extraction: Wilcoxon signed-rank test
- Reporting: median difference with IQR

All secondary analyses are interpreted descriptively unless otherwise specified. No interim analyses are planned.

### 2.8 Interpretation hierarchy

Only H1 constitutes a confirmatory primary inference. All other analyses are secondary or exploratory and are interpreted accordingly.

## 3. Study design

### 3.1 Overview

This study is designed as a prospective, protocol-driven benchmarking investigation evaluating automated extraction systems for diagnostic accuracy data under strictly controlled conditions. All critical components of the study are locked prior to execution, including the document corpus, operating procedures, data capture templates, and statistical analysis plan.

The study is intentionally conservative. Systems are evaluated exclusively on the basis of published full-text PDFs and publicly available supplementary material. External browsing, database queries, author correspondence, retrieval augmentation, and any form of adaptive prompting are explicitly prohibited during system execution. This constraint reflects the real-world conditions faced by systematic review teams at the time of extraction and prevents post hoc enrichment of incomplete reporting.

The design follows emerging methodological recommendations for the evaluation of generative artificial intelligence tools in healthcare research. In particular, the study was developed in close alignment with the Chatbot Assessment Reporting Tool (CHART) statement [10]. All relevant CHART domains, including system specification, prompt transparency, reference standard definition, failure mode characterization, reproducibility, and reporting of uncertainty, were prospectively addressed and fully implemented in the study protocol. This framework emphasizes transparency, reproducibility, and explicit definition of failure modes, including non-derivability and hallucination risk.

### 3.2 Corpus, datasets, and evaluation units

The locked corpus consists of eight peer-reviewed diagnostic accuracy studies included in a prospectively registered systematic review and meta-analysis of the Uromonitor molecular urine assay. For each publication, two diagnostic tests are queried:

1. Uromonitor
2. Urine cytology

This yields a total of 15 diagnostic datasets, reflecting the fact that urine cytology is not present in one publication. For each dataset, the atomic extraction target is a 2×2 diagnostic contingency table comprising four cells (TP, FP, FN, TN). Systems are executed repeatedly under identical conditions to allow assessment of variability and stability.

#### Evaluation units are defined as follows

- **Primary evaluation unit:** dataset-run Each dataset evaluated in a single run constitutes one binary outcome (correct or incorrect) under the end-to-end definition.
- **Secondary evaluation unit:** cell Individual TP, FP, FN, and TN cells are evaluated only for benchmark-derivable datasets.
- **Derived metrics unit:** dataset-level diagnostic metrics Sensitivity, specificity, predictive values, accuracy, and AUC_binary are derived from complete 2×2 tables where available.

This hierarchical structure avoids conflation of fundamentally different error types and aligns with prior evidence showing that apparent pointwise accuracy can mask higher-level instability.

### 3.3 Analysis sets

All datasets are classified a priori according to the human benchmark into derivable and non-derivable categories.

#### Primary derivable analysis set

Includes all datasets for which a complete 2×2 table can be unambiguously derived from the provided PDF and supplementary material according to the benchmark review. These datasets are used for evaluation of numeric extraction accuracy and downstream diagnostic metrics.

#### Safety (non-derivable) analysis set

Includes pre-specified sentinel datasets for which a complete 2×2 table is not derivable from the documents alone. These include:

- Uromonitor and urine cytology in the Rubio-Briones study [8]
- Urine cytology in the Azawi study [9]

For these datasets, the correct behavior is explicit declaration of non-derivability without numeric output. They are included in the primary end-to-end correctness endpoint as abstention cases and are additionally analyzed separately for safety and hallucination behavior. This separation ensures that abstention is treated as a clinically meaningful outcome rather than as missing data.

### 3.4 Human benchmark and reference standard

The human benchmark is defined by a rigorously conducted, prospectively registered systematic review and diagnostic accuracy meta-analysis, currently under peer review. The benchmark provides:

- An authoritative registry of included studies and datasets
- Benchmark classification of derivability based solely on document content
- Reference TP, FP, FN, and TN values for all derivable datasets
- Explicit documentation of cases where derivation is not possible without external information

For the Rubio-Briones publication, numeric contingency data were obtained by the benchmark authors through direct correspondence. Because such information is not available to automated systems restricted to published documents, these values are **not** considered derivable for the purpose of this study. Accordingly, Rubio-Briones datasets are excluded from primary numeric accuracy analyses and reserved for safety evaluation. This approach ensures that system errors are interpreted relative to a clinically realistic reference standard rather than to privileged information unavailable in routine workflows.

### 3.5 Document packages and ingestion rules

For each dataset, a frozen document package is created, consisting of:

- The full-text PDF of the publication [8,9,11–16]
- Any publicly available supplementary material

Each package is assigned a unique Input_Package_ID and provided identically to all systems.

For LLM-based systems, documents must be ingested as files. Manual copy-paste of extracted text is not permitted. If a platform fails to ingest a document fully, displays truncation warnings, or indicates partial processing, the run is classified as technically compromised, recorded as such, and excluded from consensus-level analyses while remaining visible in descriptive reporting.

These rules prevent silent context loss and ensure that observed failures reflect system behavior rather than undocumented technical limitations.

### 3.6 Systems under evaluation

#### MedNuggetizer

MedNuggetizer is executed according to its published system description, with controlled configuration and full logging [6]. It represents the primary system under validation and is evaluated across all datasets and runs.

#### Large language models

At least three contemporary LLMs are evaluated as external comparators, selected to represent distinct provider ecosystems and model architectures. For every run, the following are recorded verbatim:

- Model name and provider
- Exact model version identifier
- Access method (web interface, API, enterprise environment)
- Date and time of execution

All LLMs are run using the most deterministic settings supported by the platform, with stochastic parameters minimized and logged. Any model update during the study period is treated as a separate analytical stratum.

### 3.7 Binding standard operating procedure

System execution follows a binding SOP written as a work instruction for the study operators. Operators are responsible for:

- Executing runs exactly as specified
- Transcribing outputs verbatim into capture sheets
- Recording provenance exactly as provided by the system
- Applying derivability and safety rules strictly, without interpretation or correction

Each dataset is processed in 20 independent runs per system, each run conducted in a fresh session and temporally separated to minimize anchoring. Outputs are recorded without cleaning or reconciliation at the run level.

A random audit of at least 20% of dataset-runs is conducted by the study lead or delegate to verify transcription fidelity, provenance plausibility, and correct application of derivability rules.

### 3.8 Rationale for multi-run design

Repeated execution is not a robustness luxury but a methodological necessity. Prior work has shown that LLM-based extraction exhibits non-trivial run-to-run variability, particularly for numerically dense variables and dispersed reporting. Single-pass evaluation systematically underestimates this instability.

By enforcing 20 independent runs and explicitly analyzing repeatability, this study quantifies intraobserver variability and distinguishes between systems that are occasionally correct and systems that are reliably correct. This distinction is critical for clinical evidence synthesis, where silent variability is unacceptable.

## 4. Human benchmark and reference standard

### 4.1 Rationale for the human benchmark

A human reference standard is indispensable when evaluating automated extraction systems for diagnostic accuracy data. The benchmark must reflect how evidence synthesis is conducted in clinical research practice, including the constraints imposed by incomplete or heterogeneous reporting.

In this study, the human benchmark is derived from a prospectively registered systematic review and diagnostic accuracy meta-analysis of the Uromonitor molecular urine assay. The benchmark review was conducted according to established methodological standards for diagnostic test accuracy reviews and provides an explicit, reproducible reference for both derivability and numeric correctness.

Importantly, the benchmark distinguishes between information that is explicitly available in published documents and information that can only be obtained through external sources such as author correspondence. This distinction is critical, because automated systems evaluated in this study are deliberately restricted to published PDFs and publicly available supplementary material.

### 4.2 Benchmark derivability classification

For each diagnostic dataset, the human benchmark classifies whether a complete 2×2 contingency table (TP, FP, FN, TN) is derivable from the document package alone.

A dataset is classified as *benchmark-derivable* if and only if all four contingency table cells can be reconstructed unambiguously from the PDF and publicly available supplementary material without assumptions, inference, or back-calculation from summary statistics.

A dataset is classified as *benchmark-non-derivable* if at least one of the four cells cannot be determined with certainty from the documents alone.

This classification is performed independently of any automated system output and is fixed prior to system evaluation.

### 4.3 Reference values and handling of privileged information

For benchmark-derivable datasets, the human benchmark provides reference TP, FP, FN, and TN values that serve as the ground truth for numeric accuracy assessment.

For selected publications, including the Rubio-Briones study, additional numeric details were obtained by the benchmark authors through direct correspondence with the original investigators. While such information is valid for meta-analytic synthesis, it is not available to automated systems restricted to published documents.

Accordingly, datasets for which numeric values are only available through author correspondence are classified as non-derivable for the purposes of this study. Automated systems are not penalized for failing to reproduce privileged information that would not be accessible in routine workflows.

This approach ensures that system performance is evaluated against a clinically realistic reference standard rather than against information asymmetrically available to human reviewers.

### 4.4 Benchmark integrity and quality control

All benchmark classifications and reference values were reviewed by at least two experienced investigators with expertise in diagnostic accuracy studies. Discrepancies were resolved by consensus, with explicit documentation of the reasoning process.

The benchmark dataset and derivability labels were locked prior to initiation of system runs and were not modified during or after automated extraction.

## 5. Automated extraction systems

### 5.1 MedNuggetizer

MedNuggetizer is a confidence-based information nugget extraction system designed to support clinicians and researchers working with long-form medical documents. The system performs repeated extraction of query-relevant information nuggets, followed by clustering and consolidation to stabilize outputs and reduce omission and variability.

For the purposes of this study, MedNuggetizer is configured according to its published system description. All extraction runs are logged, and no manual post-processing, correction, or reconciliation of outputs is permitted at the run level.

MedNuggetizer represents the primary system under validation.

### 5.2 Large language models

Three contemporary large language models are evaluated as external comparators. Models are selected to represent different provider ecosystems and architectural approaches.

For each model and each run, the following metadata are recorded:

- Model name and provider
- Explicit model version identifier
- Access modality (web interface or API)
- Date and time of execution

Models are operated with the most deterministic settings supported by the platform. Stochastic parameters are minimized and documented. Any model update occurring during the study period is treated as a distinct analytical stratum.

No model-specific prompt optimization, fine-tuning, retrieval augmentation, or external browsing is permitted.

### 5.3 Prompting constraints

All systems are queried using a locked, uniform prompt, applied identically across MedNuggetizer and all LLMs. The prompt explicitly restricts extraction to absolute numbers reported in the documents and mandates explicit declaration of non-derivability when a complete 2×2 table cannot be determined with certainty. Prompt heterogeneity is intentionally avoided. This ensures that observed performance differences reflect system behavior rather than prompt engineering.

### 5.4 Prohibited system behaviors

Across all systems, the following are explicitly prohibited:

- Use of external databases, search engines, or retrieval augmentation
- Assumptions about prevalence, denominators, or cohort size
- Back-calculation from sensitivity, specificity, predictive values, or percentages unless all absolute components are explicitly reported
- Manual correction or interpretation of outputs by operators

Any violation of these rules results in the run being classified as incorrect.

## 6. System execution and standard operating procedure

### 6.1 Execution protocol

Each system is executed according to a binding standard operating procedure. For each run:

1. A fresh session is initiated.
2. The frozen document package is uploaded or ingested as a file.
3. The locked prompt is applied sequentially for Uromonitor and urine cytology.
4. Outputs are recorded verbatim.

Each system performs 20 independent runs across the full dataset. Runs are temporally separated to minimize anchoring effects.

### 6.2 Output capture and transcription

System outputs are transcribed verbatim into structured capture sheets. No normalization, correction, or reconciliation is performed at the run level.

For numeric outputs, TP, FP, FN, and TN values are recorded exactly as provided. For non-derivable outputs, the presence or absence of numeric values is explicitly documented.

Provenance information is recorded exactly as provided by the system and is not supplemented by the operators.

### 6.3 Handling of technical failures

If a system fails to ingest a document fully, produces truncation warnings, or otherwise signals incomplete processing, the run is flagged as technically compromised.

Technically compromised runs are excluded from consensus-level analyses but remain visible in descriptive reporting to ensure transparency.

### 6.4 Quality assurance and auditing

To ensure data integrity, a random sample of at least 20 % of dataset-runs is audited by the study lead or a designated delegate. Audits verify:

- Fidelity of transcription
- Correct application of derivability rules
- Plausibility and internal consistency of provenance

Any systematic deviations trigger a full review of affected runs.

### 6.5 Rationale for strict procedural control

Automated extraction systems are sensitive to small procedural deviations. By enforcing a rigid execution protocol, this study isolates system behavior from operator variability and technical confounding.

This level of control is essential for interpreting performance differences as meaningful characteristics of the systems under evaluation rather than artifacts of execution.

## 7. Data capture, data management, and audit trail

### 7.1 Data capture framework

Data capture is performed using a predefined, locked set of structured spreadsheets designed to ensure consistency, traceability, and reproducibility across all systems and runs. Four Excel-based capture sheets are mandatory and must be implemented exactly as specified prior to study initiation.

The separation of registry, run-level metadata, extraction output, and benchmark reference data is intentional. It prevents post hoc contamination of extraction outputs by benchmark information and supports transparent auditing of every extraction decision.

### 7.2 Dataset registry

The dataset registry defines the immutable structure of the evaluation corpus. For each diagnostic dataset, the registry records the publication identifier, test type, availability of supplementary material, and benchmark derivability status.

The registry is completed and locked before the first system run. No registry fields are modified during system execution. Any protocol-relevant annotations are recorded in a free-text notes field without altering core registry variables.

### 7.3 Run-level logging and traceability

Each system run is documented in a dedicated run log capturing execution metadata, including:

- unique run identifier,
- system and model version,
- access modality,
- operator,
- date and time,
- execution duration,
- and any technical issues or platform warnings.

This log provides a complete audit trail linking every extracted value to a specific system instance, configuration, and execution context. Runs flagged as technically compromised remain part of the dataset for transparency but are handled according to pre-specified rules in downstream analyses.

### 7.4 Extraction output recording

For each dataset and run, extraction outputs are recorded verbatim in the extraction output sheet. Recorded fields include:

- derivability decision,
- numeric TP, FP, FN, and TN values if derivable,
- provenance information,
- justification for non-derivability where applicable,
- and explicit flags for omissions, hallucinations, and provenance errors.

No normalization, correction, or interpretation is permitted at this stage. The extraction output sheet represents the raw system output as observed by the operator.

### 7.5 Human benchmark reference table

Benchmark reference values are stored in a separate, protected table accessible only during analysis. This table contains benchmark derivability classifications and reference TP, FP, FN, and TN values for derivable datasets.

This separation ensures that benchmark information does not influence system execution or transcription and preserves the integrity of paired comparisons during analysis.

### 7.6 Data integrity and quality assurance

To ensure data integrity, a minimum of 20 % of dataset-runs is randomly selected for audit. Audits verify transcription fidelity, plausibility of provenance statements, and correct application of derivability rules.

Audit outcomes are documented in a quality control log. Systematic errors trigger predefined corrective actions, including operator retraining or protocol amendment if necessary.

## 8. Outcome definitions and scoring

### 8.1 Primary outcome: end-to-end dataset-run correctness

The primary outcome is **end-to-end dataset-run correctness**, defined at the level of a single diagnostic dataset evaluated in a single run.

A dataset-run is classified as **correct** if and only if:

- for benchmark-derivable datasets, all four contingency table cells (TP, FP, FN, TN) exactly match the benchmark reference values; or
- for benchmark-non-derivable datasets, the system outputs “Not derivable” and outputs no numeric values.

Any deviation from these conditions results in classification as incorrect.

This binary outcome integrates numeric accuracy and correct abstention into a single, user-relevant measure of reliability.

### 8.2 Secondary outcomes: numeric extraction performance

For benchmark-derivable datasets only, numeric extraction performance is evaluated using two complementary measures:

1. Dataset-level exactness, defined as correct extraction of all four cells.
2. Cell-level exact-match accuracy, defined as the proportion of individual TP, FP, FN, and TN cells exactly matching the benchmark.

Cell-level outcomes are reported descriptively and are not used for confirmatory inference unless explicitly stated.

### 8.3 Safety and hallucination outcomes

For pre-specified benchmark-non-derivable sentinel datasets, safety is evaluated separately. Correct safety behavior is defined as explicit declaration of non-derivability without numeric output. Unsafe behavior is defined as any numeric output in this setting. Safety outcomes are reported as proportions of unsafe runs with confidence intervals and are analyzed comparatively across systems as secondary outcomes.

### 8.4 Provenance and omission errors

Provenance quality is assessed descriptively by evaluating whether systems provide plausible, document-anchored references supporting extracted values.

Omission errors are recorded when a benchmark-derivable numeric value is not provided by the system. Provenance errors are recorded when a provided reference does not plausibly support the extracted value.

These error categories inform qualitative interpretation of system behavior and are not used for confirmatory hypothesis testing.

### 8.5 Derived diagnostic metrics

For benchmark-derivable datasets with complete contingency tables, diagnostic performance metrics are derived using standard definitions:

- sensitivity,
- specificity,
- positive predictive value,
- negative predictive value,
- accuracy.

To ensure comparability across datasets and systems, a prespecified binary AUC is calculated as the mean of sensitivity and specificity. This metric reflects performance at the single reported operating point and avoids assumptions inherent to continuous-score ROC reconstruction. An exploratory F1 score is calculated as the harmonic mean of precision and sensitivity and is reported descriptively.

## 9. Multi-run handling, consensus rules, and repeatability

### 9.1 Rationale for multi-run evaluation

Single-pass evaluation of automated extraction systems underestimates variability and masks instability that is clinically relevant. Repeated execution allows explicit quantification of intraobserver variability and supports cautious real-world use through consensus formation.

Accordingly, each system is evaluated across 20 independent runs per dataset.

### 9.2 Run-level and consensus-level reporting

Two complementary reporting layers are defined. The run-level layer captures raw variability, error modes, and stochastic behavior across individual runs and constitutes the primary level for confirmatory statistical inference, including non-inferiority testing.

The consensus-level layer reflects cautious real-world use, in which repeated extractions are reconciled to improve robustness. Consensus-level results are reported as supportive robustness analyses and to illustrate practical application under repeated-use conditions, and are not used for confirmatory hypothesis testing.

### 9.3 Consensus derivability rule

For each dataset and system, derivability status at the consensus level is determined by majority rule:

- classified as derivable if at least 11 of 20 runs output derivable;
- classified as non-derivable if at least 11 of 20 runs output non-derivable.

### 9.4 Consensus numeric rule

For consensus-derivable datasets, numeric values for TP, FP, FN, and TN are determined by taking the median of the values provided across runs.

### 9.5 Safety override rule

To prevent unstable or sparsely supported numeric outputs from driving consensus results, a safety override is applied. If fewer than 11 runs provide a numeric value for any contingency table cell, the dataset is classified as non-derivable at the consensus level, regardless of the derivability majority vote.

### 9.6 Assessment of intraobserver variability

Repeatability is evaluated separately for Uromonitor and urine cytology.

Derivability repeatability is quantified using Fleiss kappa across runs for each dataset, supplemented by % agreement.

Numeric repeatability for benchmark-derivable datasets is assessed by:

- the proportion of runs yielding identical complete contingency tables, and
- intraclass correlation coefficients for individual cells and derived metrics.

Repeatability outcomes are summarized per system and compared descriptively and inferentially as specified in the statistical analysis plan.

### 9.7 Interpretation of repeatability results

High repeatability indicates that correct behavior is stable and reproducible rather than incidental. Low repeatability, even with acceptable average accuracy, is interpreted as a limitation for clinical evidence synthesis, where consistency is essential.

## 10. Statistical analysis plan

### 10.1 General principles

All statistical analyses will be conducted using IBM SPSS Statistics, version 31.0.

Unless explicitly stated otherwise, tests are two-sided. Non-inferiority testing is performed one-sided, as prespecified.

Confirmatory inference is limited to the primary non-inferiority hypothesis for MedNuggetizer. All other hypotheses are secondary or exploratory and are interpreted accordingly, with multiplicity handling as specified.

Analyses are performed on non-compromised runs only. Runs flagged as technically compromised remain visible in descriptive reporting but are excluded from inferential analyses and from consensus derivations.

### 10.2 Analysis populations and statistical units Run-level population

Includes all dataset-runs without technical compromise.

#### Consensus-level population

Derived from run-level data using the consensus rules defined in Chapter 9.

#### Sentinel safety population

Includes only the pre-specified non-derivable datasets (Azawi urine cytology; Rubio-Briones Uromonitor and urine cytology).

#### Primary statistical unit

Dataset-run, classified as correct or incorrect under the end-to-end definition.

#### Secondary units

- Dataset (for repeatability and consensus analyses)
- Cell (TP, FP, FN, TN) for descriptive numeric accuracy
- Run (for time analyses)

### 10.3 Variable derivation for analysis

The following variables are derived directly from the capture sheets:

#### Primary correctness variable

Dataset-run correctness is coded as binary (1 = correct, 0 = incorrect), according to the definition in Chapter 8.

#### Hallucination variable

For sentinel non-derivable datasets, a hallucination event is coded as 1 if any numeric TP, FP, FN, or TN value is output, and 0 if the output is “Not derivable” without numeric values.

#### Repeatability category variable

Each dataset-run is classified into one of three mutually exclusive categories:

1. Correct numeric extraction
2. Correct abstention
3. Incorrect or unsafe output

#### Execution time variable

Run duration in seconds, defined as the interval from run initiation to completion of protocol-conform output for the full corpus.

### 10.4 Primary endpoint analysis (H1): non-inferiority of MedNuggetizer

#### 10.4.1 Hypothesis and estimand

Let p denote the probability that MedNuggetizer produces a correct end-to-end output for a dataset-run.

Null hypothesis: p ≤ 0.95

Alternative hypothesis: p > 0.95

The non-inferiority margin is therefore defined as an absolute threshold of 95 % correctness. The test is one-sided with alpha equal to 0.025. Primary inference is based on run-level dataset-run correctness. Consensus-level results derived from repeated runs are reported as supportive robustness analyses and are not used for confirmatory hypothesis testing.

#### 10.4.2 Statistical test and decision rule

The primary analysis uses an exact binomial framework.

For MedNuggetizer:

- Let N be the total number of non-compromised dataset-runs.
- Let X be the number of correct dataset-runs.

The primary decision criterion is based on the exact one-sided 97.5 % lower confidence bound for p using the Clopper–Pearson method.

Non-inferiority is declared if and only if the lower confidence bound exceeds 0.95.

In addition, the one-sided exact p-value for the test p > 0.95 is reported.

#### 10.4.3 SPSS 31.0 implementation

In SPSS:

1. Filter the dataset to include only MedNuggetizer runs without technical issues.
2. Use descriptive statistics or frequencies to obtain X (sum of correct runs) and N (total runs).
3. The exact one-sided confidence bound and one-sided p-value are calculated using the exact binomial distribution functions available in SPSS.
4. The resulting lower bound is compared against the fixed threshold of 0.95 to determine non-inferiority.

The observed proportion X/N, the lower confidence bound, and the one-sided p-value are all reported.

### 10.5 Secondary non-inferiority analyses (H2): LLMs

Each evaluated LLM is tested analogously to MedNuggetizer using the same exact binomial framework and the same non-inferiority threshold of 95 % correctness.

Multiplicity across the three LLM non-inferiority tests is handled using the Holm procedure with a family-wise alpha of 0.025.

For each LLM, the following are reported:

- Observed accuracy (X/N)
- One-sided 97.5 % lower exact confidence bound
- One-sided exact p-value
- Holm-adjusted non-inferiority decision

### 10.6 Safety and hallucination analysis (H3)

Safety analyses are restricted to the sentinel non-derivable datasets. For each system:

- The hallucination rate is calculated as the proportion of sentinel dataset-runs with any numeric output.
- Exact confidence intervals for the hallucination rate are computed using the Clopper–Pearson method.

Between-system comparisons are performed using Fisher’s exact test, comparing MedNuggetizer against each LLM separately.

Multiplicity across these comparisons is handled using the Holm procedure with a family-wise alpha of 0.05, as this analysis is secondary.

SPSS implementation uses crosstabulation with exact tests enabled.

### 10.7 Repeatability and intraobserver variability (H4)

Repeatability is assessed at two levels.

#### 10.7.1 Output-category repeatability

For each dataset and system, agreement across the 20 runs for the three-category output variable is quantified using Fleiss’ kappa.

For each system:

- Fleiss’ kappa is computed per dataset.
- Median and interquartile range of kappa values are reported across datasets.

Between-system comparisons of repeatability are performed using dataset-paired Wilcoxon signed-rank tests comparing MedNuggetizer with each LLM. Holm adjustment is applied across the three comparisons.

#### 10.7.2 Numeric repeatability for derivable datasets

For benchmark-derivable datasets:

- The most frequent complete 2×2 table across runs is identified for each dataset and system.
- Numeric stability is defined as the proportion of runs that exactly match this modal table.

Numeric stability proportions are summarized per system using median and interquartile range.

Between-system comparisons use dataset-paired Wilcoxon signed-rank tests with Holm adjustment.

### 10.8 Operational efficiency analysis (H5)

Execution time is summarized descriptively for each system using median, interquartile range, minimum, and maximum.

Formal comparisons against human extraction time are performed using paired Wilcoxon signed-rank tests, pairing runs by run index, as run timing is captured consistently across systems.

Technically compromised runs are excluded from inferential time analyses.

### 10.9 Additional descriptive analyses

The following analyses are descriptive and not used for confirmatory inference:

- Cell-level exact-match accuracy for TP, FP, FN, and TN with exact confidence intervals.
- Rates of omission errors and provenance errors by system and test type.
- Absolute and signed errors for derived diagnostic metrics (sensitivity, specificity, PPV, NPV, accuracy, AUC_binary), summarized by median and interquartile range.
- Exploratory F1 scores.

### 10.10 Reporting standards and reproducibility

All reported results will include the corresponding denominators, confidence intervals, and exact test specifications. Decisions regarding non-inferiority, safety, and repeatability will be traceable to prespecified thresholds and statistical tests. The full statistical analysis follows a locked analysis plan and is reproducible using the captured run logs, extraction outputs, and benchmark reference tables.

## 11. Manuscript blueprint for the primary publication

### 11.1 Core message

This study provides a rigorous, clinically grounded answer to a central question in modern urologic evidence synthesis: whether automated systems, including large language models and a purpose-built extraction pipeline, can be used reliably and safely to extract diagnostic accuracy data from full-text publications.

The manuscript explicitly addresses four dimensions that are critical for real-world use: end-to-end correctness, repeatability across repeated runs, safety in non-derivable reporting scenarios, and downstream consequences for clinically relevant diagnostic performance metrics. The emphasis is not on maximal output, but on reproducible and trustworthy behavior under realistic constraints.

### 11.2 Planned tables and figures

The manuscript will include the following prespecified tables and figures:

- **Table 1.** Corpus characteristics and benchmark derivability classification
- **Table 2.** End-to-end dataset-level correctness and numeric extraction performance
- **Table 3.** Error taxonomy by system, including omissions, hallucinations, and provenance errors
- **Table 4.** Repeatability across runs, including Fleiss kappa and numeric stability metrics
- **Table 5.** Fidelity of derived diagnostic performance metrics for Uromonitor and urine cytology
- **Figure 1.** Study workflow and evaluation framework
- **Figure 2.** Distribution of absolute errors in diagnostic metrics by system and test type represents the primary system Safety behavior on pre-specified non-derivable sentinel datasets

All tables and figures are designed to be interpretable without reference to supplementary material.

### 11.3 Practical recommendations for evidence synthesis teams

A dedicated section of the Discussion will translate quantitative findings into operational guidance for urologic systematic review teams.

Based on the study design, this section will articulate minimum standards for responsible use of automated extraction systems, including:

- execution of multiple independent runs rather than single-pass extraction,
- mandatory documentation of provenance,
- explicit declaration of non-derivability when source documents are insufficient,
- and use of conservative consensus rules to mitigate run-to-run variability.

These recommendations are intended to support informed, transparent integration of automated tools into evidence synthesis workflows rather than to promote unsupervised automation.

### 11.4 Generalizability beyond urologic diagnostics

Although the benchmark corpus is grounded in uro-oncologic diagnostic accuracy studies, the methodological framework developed in this study is not disease- or specialty-specific. The principles of explicit derivability classification, end-to-end correctness assessment, multi-run repeatability analysis, and evaluation of downstream effects on diagnostic performance metrics are directly transferable to diagnostic systematic reviews in other medical disciplines. The Discussion will explicitly address how these safeguards may inform safe and reproducible use of automated extraction systems beyond urology.

## 12. Target journals and publication strategy

This study is positioned as a clinically anchored methodological contribution rather than a pure informatics benchmark. Target journals are selected accordingly.

### Primary target journals

- *European Urology Focus:* Strong alignment with the urologic readership and a clear track record of publishing clinically relevant methodological and translational studies. The journal is receptive to work that bridges clinical practice and emerging technologies, particularly when grounded in realistic workflows and rigorous methodology. The present study fits well within the scope of evidence synthesis, diagnostics, and responsible AI use in urology.
- *BJU International:* Well-established international urology journal with a strong interest in methodological rigor, diagnostics, and innovation that directly impacts clinical decision-making. The journal frequently publishes studies on diagnostic pathways, biomarkers, and evidence synthesis, making it a realistic and credible target.

### Secondary target journal

- *Journal of the American Medical Informatics Association (JAMIA):* Strong methodological alignment, particularly for the extraction and repeatability components, with the requirement that generalizability beyond the Uromonitor use case is clearly articulated.
- *Artificial Intelligence in Medicine:* Longstanding journal at the interface of clinical medicine and AI, with a tradition of publishing rigorous methodological evaluations rather than purely performance-driven benchmarks. The conservative, safety-aware design of this study aligns well with the journal’s scope.

Journal fit scores reflect scope alignment, readership relevance, and anticipated receptiveness to a rigorously designed methods study embedded in a clinical context.

## 13. Ethics, transparency, and reproducibility

This study does not involve patient-level data, human participants, or interventions. All inputs consist of published full-text articles and publicly available supplementary material. Accordingly, institutional review board approval was not required.

To promote transparency and reproducibility, run logs, capture templates, consensus rules, and analysis definitions will be made available as supplementary material where permitted by journal policy. This will allow independent replication and reanalysis of all reported results.

### 13.1 AI reporting framework and governance

The design, conduct, and reporting of this study are informed by the Chatbot Assessment Reporting Tool (CHART) statement, a consensus-based reporting guideline for evaluation of generative artificial intelligence systems in healthcare [10].

In alignment with CHART principles, this protocol prepecifies model identifiers and versions, access routes, prompting strategy, evaluation metrics, and a human reference standard.

Safety-relevant failure modes, including non-derivable outputs, hallucinations, omissions, and provenance errors, are defined prospectively. Reproducibility is addressed through repeated independent runs and explicit consensus rules.

Automated systems are treated as assistive research tools rather than autonomous decision-makers. All outputs are audited against a locked benchmark under a predefined statistical analysis plan to ensure interpretability and accountability.

## 14. Selection of large language models for the benchmark

### 14.1 Rationale for a three-model design

In addition to MedNuggetizer [6], three contemporary large language models are evaluated. The three-model design is intentional. It limits multiplicity, preserves interpretability in paired comparisons, and reflects the practical reality of academic urology groups, which typically evaluate a small number of tools for routine use.

The objective is not a market survey, but a clinically actionable comparison under controlled conditions.

### 14.2 Model selection criteria

Each LLM included in the benchmark must meet the following criteria:

1. Demonstrated capability for document-based numeric extraction from full-text PDFs.
2. Robust handling of long and heterogeneous documents, including tables and figure-associated text.
3. Mature document ingestion functionality for PDFs and supplements.
4. Ability to record and lock explicit model identifiers and versions.
5. Realistic availability to academic clinical research teams.

### 14.3 OpenAI GPT-5.2

GPT-5.2 is included as a representative of the OpenAI GPT model family widely used in academic and clinical research settings [17]. Its documented support for versioned access enables precise audit trails.

The exact model identifier used for each run will be recorded verbatim. Any provider-side update during the study window will be treated as a separate analytical stratum.

### 14.4 Anthropic Claude Opus 4.5

Claude Opus 4.5 represents a distinct model lineage and provider ecosystem, strengthening external validity [18]. Its inclusion is methodologically relevant for the safety component of this study, as the benchmark includes pre-specified non-derivable scenarios that require explicit abstention rather than numeric guessing.

Model identifiers are recorded and locked per run. Any change in model designation during execution is handled as protocol-defined model drift.

### 14.5 Google Gemini 3 Pro

Gemini 3 Pro provides a third independent model family with strong multimodal document understanding, relevant for diagnostic studies in which key values may appear in tables, captions, or figure annotations [19].

As with the other models, the exact identifier is captured for every run. Platform messages indicating substitution or silent upgrades are documented and handled according to the protocol.

### 14.6 Configuration lock and documentation

For every LLM run, the following parameters are documented:

- provider and access modality,
- exact model identifier and version,
- date and time of execution,
- document ingestion method,
- decoding parameters and reproducibility settings,
- confirmation that external tools, browsing, and retrieval are disabled,
- and any truncation or partial-processing warnings.

These requirements ensure that observed variability reflects intrinsic system behavior rather than uncontrolled platform features.

### 14.7 Justification for limiting the benchmark to three LLMs

Expanding the benchmark beyond three LLMs would increase multiplicity and dilute interpretability without proportionate gain in insight, given the paired design and fixed corpus size.

The selected models are sufficient to address the central clinical question: whether a small set of widely accessible, frontier LLMs can be used reliably and safely for diagnostic data extraction in comparison with a human benchmark and a purpose-built extraction system.

### 14.8 Rationale for exclusion of DeepSeek models

DeepSeek models were considered during the planning phase of this benchmark but were not included in the final study design [20]. The primary reason for exclusion was the absence, at the time of protocol finalization, of a sufficiently stable and transparent documentation framework that would allow prospective locking of exact model identifiers, version history, and update behavior comparable to the other evaluated systems. For this study, explicit versioning, auditable model identifiers, and clearly documented update policies were mandatory prerequisites, as they directly affect reproducibility and interpretability of repeated runs. In addition, access pathways for DeepSeek models were heterogeneous across platforms, with limited standardization of document ingestion workflows for long-form PDFs and supplementary material under controlled, browsing-disabled conditions. This limited the ability to guarantee consistent execution conditions across repeated runs and across systems. Importantly, exclusion of DeepSeek models does not imply inferior performance or capability. Rather, it reflects a deliberate methodological decision to prioritize systems for which version control, access modality, and operational constraints could be prospectively locked and uniformly enforced throughout the study period. Future benchmark iterations may include DeepSeek models once stable, versioned documentation and controlled execution pathways comparable to those required by this protocol become available.

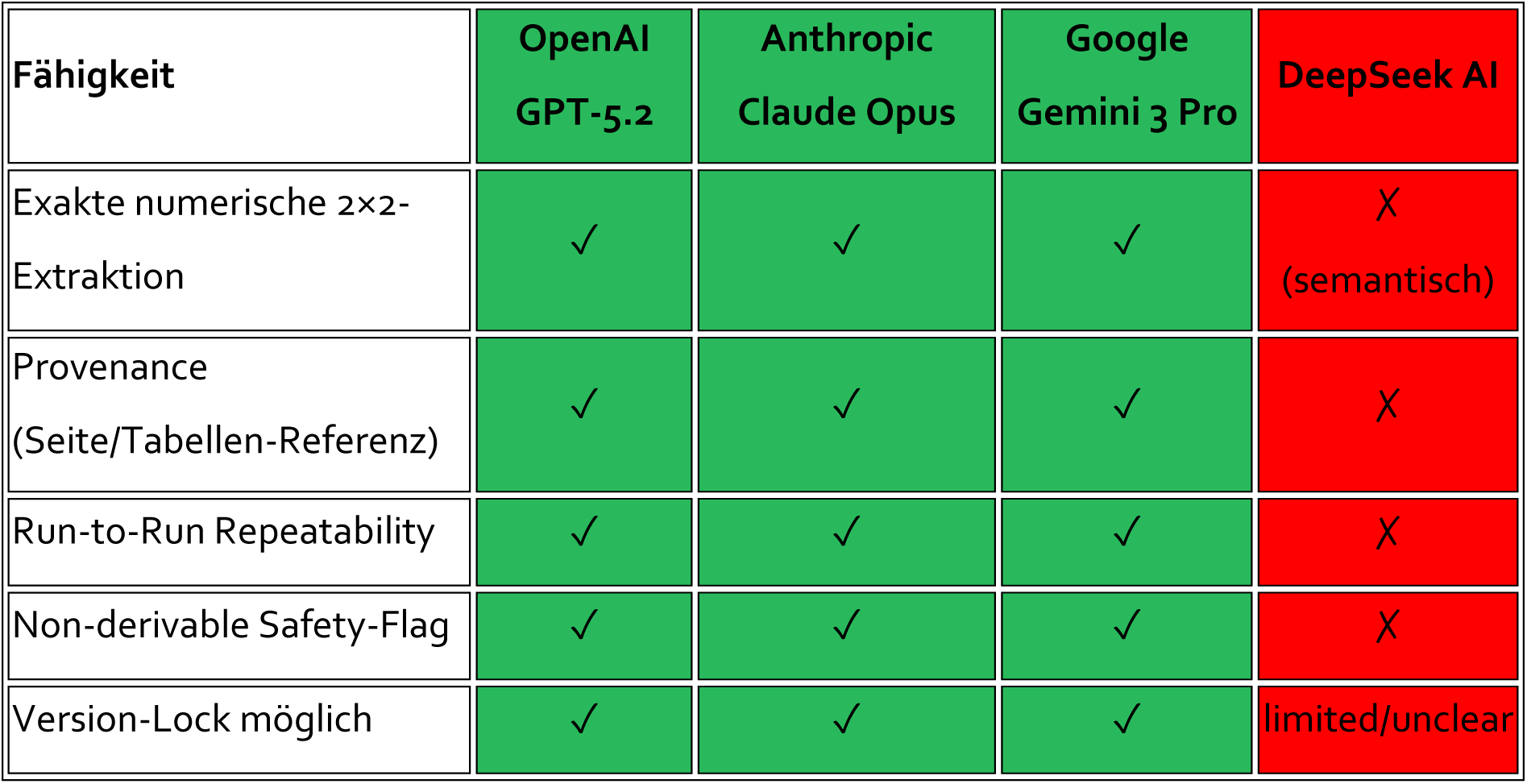

| Fähigkeit | OpenAI<br>GPT-5.2 | Anthropic<br>Claude Opus | Google<br>Gemini 3 Pro | DeepSeek AI |
| --- | --- | --- | --- | --- |
| Exakte numerische 2×2-Extraktion | ✓ | ✓ | ✓ | ✗<br>(semantisch) |
| Provenance<br>(Seite/Tabellen-Referenz) | ✓ | ✓ | ✓ | ✗ |
| Run-to-Run Repeatability | ✓ | ✓ | ✓ | ✗ |
| Non-derivable Safety-Flag | ✓ | ✓ | ✓ | ✗ |
| Version-Lock möglich | ✓ | ✓ | ✓ | limited/unclear |

## 15. Canonical extraction prompt (locked version)

**Prompt ID:** Canonical_Extraction_Prompt_v1.0

**Scope:** Diagnostic accuracy data extraction

**Target variables:** True Positives (TP), False Positives (FP), False Negatives (FN), True Negatives (TN)

**Permitted inputs:** Provided full-text PDF(s) and publicly available supplementary material only

**External sources:** Not permitted

### Uniform canonical extraction prompts

The following prompts are applied **identically** across all evaluated systems, including MedNuggetizer and all large language models.

No system-specific prompt modification, optimization, or augmentation is permitted.

### Prompt A: Uromonitor

What are the true-positive, false-positive, false-negative, and true-negative test results of the Uromonitor test in the detection of bladder cancer.

1. Use only explicitly reported absolute numbers.
2. Do not reconstruct values from sensitivity, specificity, positive predictive value, negative predictive value, accuracy, or prevalence unless all required absolute components are explicitly stated in the document.
3. If any cell of the 2×2 table cannot be determined with certainty, output “Not derivable” for the entire table.

### Prompt B: Urine cytology

What are the true-positive, false-positive, false-negative, and true-negative test results of urine cytology in the detection of bladder cancer.

1. Use only explicitly reported absolute numbers.
2. Do not reconstruct values from sensitivity, specificity, positive predictive value, negative predictive value, accuracy, or prevalence unless all required absolute components are explicitly stated in the document.
3. If any cell of the 2×2 table cannot be determined with certainty, output “Not derivable” for the entire table.

### Extraction rules and interpretation

Extraction is strictly limited to information contained in the provided full-text PDF and its publicly available supplementary material.

The following actions are explicitly prohibited:

- assumptions regarding prevalence, denominators, or cohort size,
- estimations or approximations,
- back-calculation from percentages or summary statistics,
- use of external knowledge, prior training data, or general medical knowledge.

If one or more of the four diagnostic cells (TP, FP, FN, TN) cannot be derived **with certainty** from the provided documents alone, the correct output is **“Not derivable.”**

Correct identification of non-derivability is considered a **correct and preferred outcome** and takes precedence over producing unsupported numeric values.

### Required output structure

Each run must return **exactly one** of the following outputs.

### If derivable

- True positives (TP): integer
- False positives (FP): integer
- False negatives (FN): integer
- True negatives (TN): integer
- Provenance for each value, specified as page number(s) and table, figure, or explicit text reference(s)

### If not derivable

- Output: Not derivable
- Brief justification identifying which value(s) are missing or ambiguous and why a complete 2×2 table cannot be derived

No additional commentary, interpretation, or explanation is permitted outside this structure.

### Protocol-level binding

This canonical extraction prompt is **locked** for the duration of the study. Any deviation from the prompt text, output structure, or extraction rules constitutes a protocol deviation and requires a formal protocol amendment prior to further analysis.

## Data Availability

All data produced in the present study are available upon reasonable request to the authors.

## 17. Methodological Context of Prior Evidence Informing the Present Study

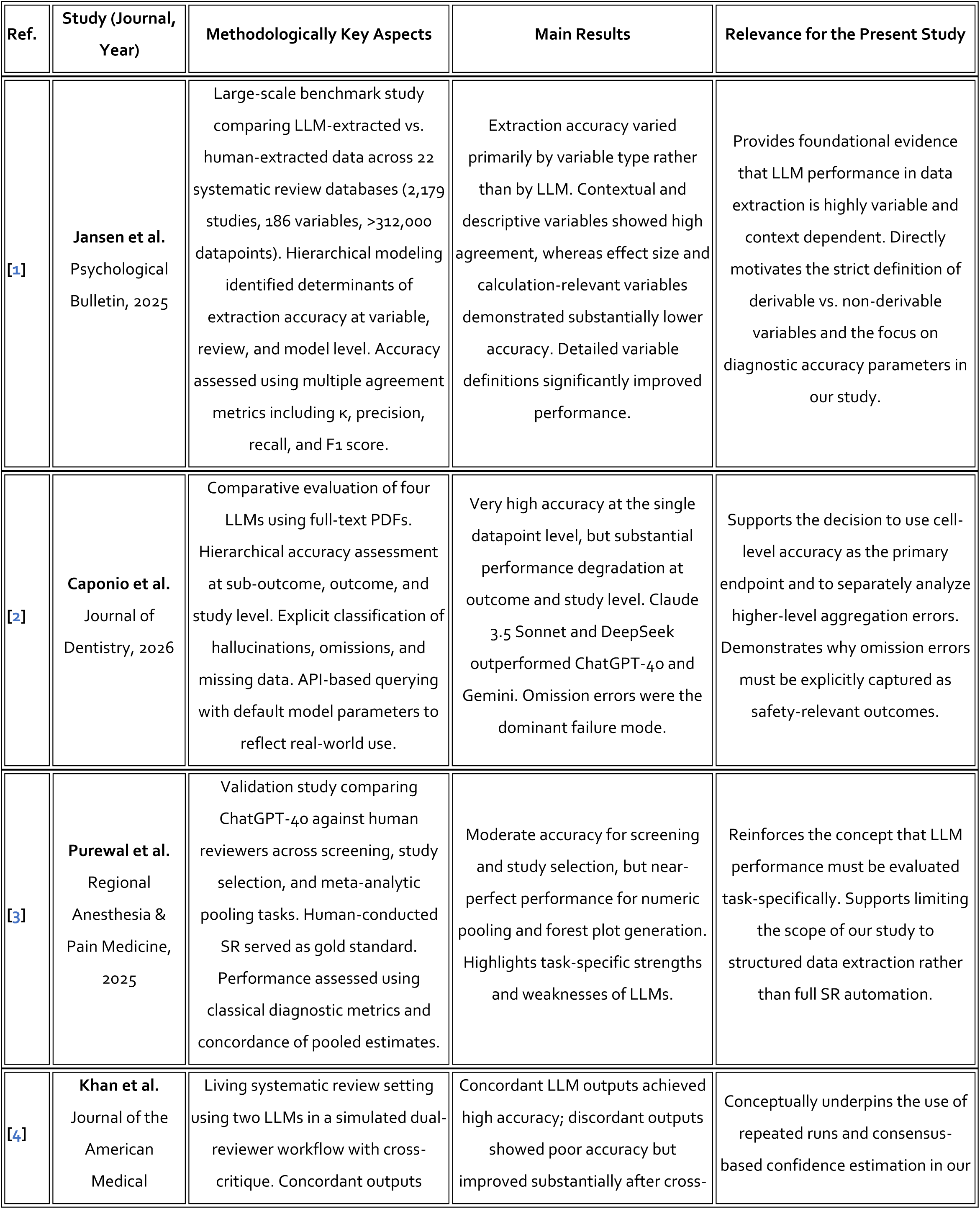
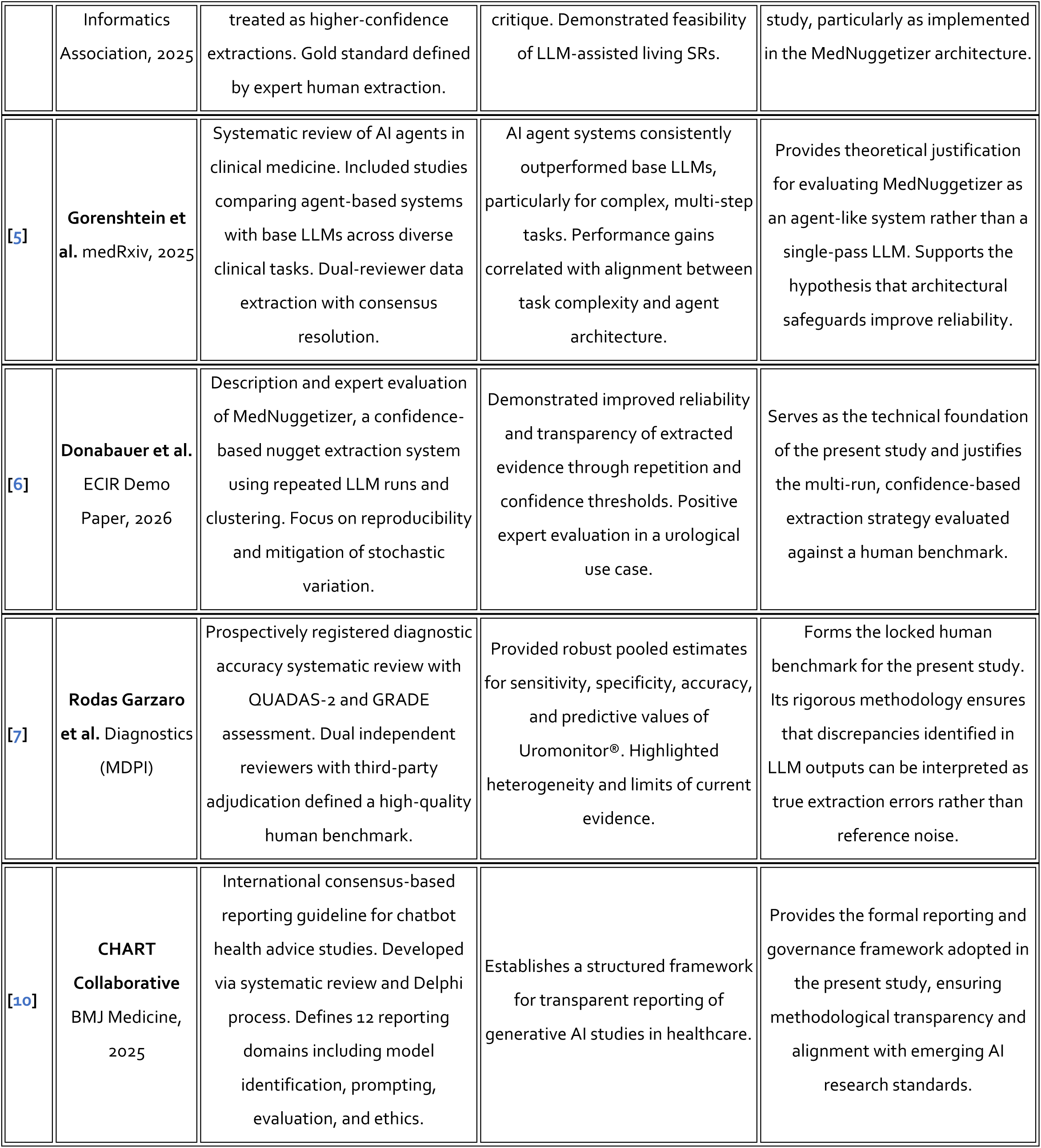

| Ref. | Study (Journal, Year) | Methodologically Key Aspects | Main Results | Relevance for the Present Study |
| --- | --- | --- | --- | --- |
| [1] | Jansen et al.<br>Psychological Bulletin, 2025 | Large-scale benchmark study comparing LLM-extracted vs. human-extracted data across 22 systematic review databases (2,179 studies, 186 variables, >312,000 datapoints). Hierarchical modeling identified determinants of extraction accuracy at variable, review, and model level. Accuracy assessed using multiple agreement metrics including $\kappa$, precision, recall, and F1 score. | Extraction accuracy varied primarily by variable type rather than by LLM. Contextual and descriptive variables showed high agreement, whereas effect size and calculation-relevant variables demonstrated substantially lower accuracy. Detailed variable definitions significantly improved performance. | Provides foundational evidence that LLM performance in data extraction is highly variable and context dependent. Directly motivates the strict definition of derivable vs. non-derivable variables and the focus on diagnostic accuracy parameters in our study. |
| [2] | Caponio et al.<br>Journal of Dentistry, 2026 | Comparative evaluation of four LLMs using full-text PDFs. Hierarchical accuracy assessment at sub-outcome, outcome, and study level. Explicit classification of hallucinations, omissions, and missing data. API-based querying with default model parameters to reflect real-world use. | Very high accuracy at the single datapoint level, but substantial performance degradation at outcome and study level. Claude 3.5 Sonnet and DeepSeek outperformed ChatGPT-4o and Gemini. Omission errors were the dominant failure mode. | Supports the decision to use cell-level accuracy as the primary endpoint and to separately analyze higher-level aggregation errors. Demonstrates why omission errors must be explicitly captured as safety-relevant outcomes. |
| [3] | Purewal et al.<br>Regional Anesthesia & Pain Medicine, 2025 | Validation study comparing ChatGPT-4o against human reviewers across screening, study selection, and meta-analytic pooling tasks. Human-conducted SR served as gold standard. Performance assessed using classical diagnostic metrics and concordance of pooled estimates. | Moderate accuracy for screening and study selection, but near-perfect performance for numeric pooling and forest plot generation. Highlights task-specific strengths and weaknesses of LLMs. | Reinforces the concept that LLM performance must be evaluated task-specifically. Supports limiting the scope of our study to structured data extraction rather than full SR automation. |
| [4] | Khan et al.<br>Journal of the American Medical | Living systematic review setting using two LLMs in a simulated dual-reviewer workflow with cross-critique. Concordant outputs | Concordant LLM outputs achieved high accuracy; discordant outputs showed poor accuracy but improved substantially after cross- | Conceptually underpins the use of repeated runs and consensus-based confidence estimation in our |
|  | Informatics Association, 2025 | treated as higher-confidence extractions. Gold standard defined by expert human extraction. | critique. Demonstrated feasibility of LLM-assisted living SRs. | study, particularly as implemented in the MedNuggetizer architecture. |
| [5] | Gorenshtein et al. medRxiv, 2025 | Systematic review of AI agents in clinical medicine. Included studies comparing agent-based systems with base LLMs across diverse clinical tasks. Dual-reviewer data extraction with consensus resolution. | AI agent systems consistently outperformed base LLMs, particularly for complex, multi-step tasks. Performance gains correlated with alignment between task complexity and agent architecture. | Provides theoretical justification for evaluating MedNuggetizer as an agent-like system rather than a single-pass LLM. Supports the hypothesis that architectural safeguards improve reliability. |
| [6] | Donabauer et al. ECIR Demo Paper, 2026 | Description and expert evaluation of MedNuggetizer, a confidence-based nugget extraction system using repeated LLM runs and clustering. Focus on reproducibility and mitigation of stochastic variation. | Demonstrated improved reliability and transparency of extracted evidence through repetition and confidence thresholds. Positive expert evaluation in a urological use case. | Serves as the technical foundation of the present study and justifies the multi-run, confidence-based extraction strategy evaluated against a human benchmark. |
| [7] | Rodas Garzaro et al. Diagnostics (MDPI) | Prospectively registered diagnostic accuracy systematic review with QUADAS-2 and GRADE assessment. Dual independent reviewers with third-party adjudication defined a high-quality human benchmark. | Provided robust pooled estimates for sensitivity, specificity, accuracy, and predictive values of Uromonitor®. Highlighted heterogeneity and limits of current evidence. | Forms the locked human benchmark for the present study. Its rigorous methodology ensures that discrepancies identified in LLM outputs can be interpreted as true extraction errors rather than reference noise. |
| [10] | CHART Collaborative BMJ Medicine, 2025 | International consensus-based reporting guideline for chatbot health advice studies. Developed via systematic review and Delphi process. Defines 12 reporting domains including model identification, prompting, evaluation, and ethics. | Establishes a structured framework for transparent reporting of generative AI studies in healthcare. | Provides the formal reporting and governance framework adopted in the present study, ensuring methodological transparency and alignment with emerging AI research standards. |

## Notes

**Conflict of Interest Statement:** The authors declare no conflicts of interest. Members of the study team were involved in the academic development of the MedNuggetizer system. However, there are no financial, commercial, or proprietary interests related to MedNuggetizer, and no author has received remuneration, equity, or other financial benefit in connection with its development or evaluation.

### Competing Interest Statement

The authors declare no conflicts of interest. Members of the study team were involved in the academic development of the MedNuggetizer system. However, there are no financial, commercial, or proprietary interests related to MedNuggetizer, and no author has received remuneration, equity, or other financial benefit in connection with its development or evaluation.

### Funding Statement

This study did not receive any funding.

## References

1. Jansen T, Liebenow LW, Mertens U, Schmidt FTC, Lohmann JF, Fleckenstein J, Meyer J. Data extraction by generative artificial intelligence: Assessing determinants of accuracy using human-extracted data from systematic review databases. Psychol Bull. 2025 Oct;151(10):1280–1306. doi: 10.1037/bul0000501. PMID: 41396533.

2. Caponio VCA, Lorenzo-Pouso AI, Magalhaes M, Ali A, Adamo D, Cirillo N, López-Pintor RM, Musella G. Accuracy of LLMs to retrieve numeric data for meta-analysis in dentistry. J Dent. 2026 Jan;164:106245. doi: 10.1016/j.jdent.2025.106245. Epub 2025 Nov 19. PMID: 41265689.

3. Purewal A, Fautsch K, Klasova J, Hussain N, D’Souza RS. Human versus artificial intelligence: evaluating ChatGPT’s performance in conducting published systematic reviews with meta-analysis in chronic pain research. Reg Anesth Pain Med. 2025 Feb 16:rapm-2024-106358. doi: 10.1136/rapm-2024-106358. Epub ahead of print. PMID: 39956557.

4. Khan MA, Ayub U, Naqvi SAA, Khakwani KZR, Sipra ZBR, Raina A, Zhou S, He H, Saeidi A, Hasan B, Rumble RB, Bitterman DS, Warner JL, Zou J, Tevaarwerk AJ, Leventakos K, Kehl KL, Palmer JM, Murad MH, Baral C, Riaz IB. Collaborative large language models for automated data extraction in living systematic reviews. J Am Med Inform Assoc. 2025 Apr 1;32(4):638–647. doi: 10.1093/jamia/ocae325. PMID: 39836495; PMCID: PMC12005628.doi:10.1093/jamia/ocae325.

5. Gorenshtein A, Omar M, Glicksberg BS, Nadkarni GN, Klang E. AI Agents in Clinical Medicine: A Systematic Review. medRxiv [Preprint]. 2025 Aug 26:2025.08.22.25334232. doi: 10.1101/2025.08.22.25334232. PMID: 40909853; PMCID: PMC12407621.

6. Donabauer G, Ateia S, Kruschwitz U, Burger M, May M, Haas M, Rodas Garzaro JR, Eckl C. MedNuggetizer: Confidence-Based Information Nugget Extraction from Medical Documents. ECIR 2026 Demo Paper.

7. Rodas Garzaro JR, Kravchuk A, Wolff I, Lebentrau S, Rubio-Briones J, Brás JP, Gilfrich C, Siepmann S, Pahernik S, Merseburger A, Heidenreich A, May M. Diagnostic Performance and Clinical Utility of the Uromonitor® Molecular Urine Assay for Urothelial Carcinoma of the Bladder: A Systematic Review and Diagnostic Accuracy Meta-analysis. Diagnostics (MDPI). (diagnostics-4082381).

8. Rubio-Briones J, Guerrero Ramos F, Mercadé Sánchez A, Bezana Abadía I, Rodríguez RM, Alcaraz A, Miqueleiz Legaz M, Lozano F, Arrabal Martín M, Luis Cardo A, Gonzalo Rodríguez V, Caño Velasco J, Serrano Liesa M, Ferreiro Pareja C, Espílez Ortiz R, de González-Valcárcel de Torres I, Sánchez Zalabardo D, Esquinas C, Mengual L, Borque-Fernando A, Esteban Escaño LM, Palou Redorta J, Witjes F, Martínez Piñeiro L. External validation of the Uromonitor®-version 2 urine test as a biomarker for optimisation of non-muscle-invasive bladder cancer management. BJU Int. 2026 Jan;137(1):216–224. doi: 10.1111/bju.70010. Epub 2025 Oct 1. PMID: 41031577.

9. Azawi N, Vásquez JL, Dreyer T, Guldhammer CS, Saber Al-Juboori RM, Nielsen AM, Jensen JB. Surveillance of Low-Grade Non-Muscle Invasive Bladder Tumors Using Uromonitor: SOLUSION Trial. Cancers (Basel). 2023 Apr 17;15(8):2341. doi: 10.3390/cancers15082341. PMID: 37190269; PMCID: PMC10137147.

10. CHART Collaborative. Reporting guideline for chatbot health advice studies: the Chatbot Assessment Reporting Tool (CHART) statement. BMJ Med. 2025 Aug 1;4(1):e001632. doi: 10.1136/bmjmed-2025-001632. PMID: 40761518; PMCID: PMC12320030.

11. Batista R, Vinagre J, Prazeres H, Sampaio C, Peralta P, Conceição P, Sismeiro A, Leão R, Gomes A, Furriel F, Oliveira C, Torres JN, Eufrásio P, Azinhais P, Almeida F, Gonzalez ER, Bidovanets B, Ecke T, Stinjs P, Pascual ÁS, Abdelmalek R, Villafruela A, Beardo-Villar P, Fidalgo N, Öztürk H, Gonzalez-Enguita C, Monzo J, Lopes T, Álvarez-Maestro M, Servan PP, De La Cruz SMP, Perez MPS, Máximo V, Soares P. Validation of a Novel, Sensitive, and Specific Urine-Based Test for Recurrence Surveillance of Patients With Non-Muscle-Invasive Bladder Cancer in a Comprehensive Multicenter Study. Front Genet. 2019 Dec 18;10:1237. doi: 10.3389/fgene.2019.01237. PMID: 31921291; PMCID: PMC6930177.

12. Sieverink CA, Batista RPM, Prazeres HJM, Vinagre J, Sampaio C, Leão RR, Máximo V, Witjes JA, Soares P. Clinical Validation of a Urine Test (Uromonitor-V2®) for the Surveillance of Non-Muscle-Invasive Bladder Cancer Patients. Diagnostics (Basel). 2020 Sep 24;10(10):745. doi: 10.3390/diagnostics10100745. PMID: 32987933; PMCID: PMC7599569.

13. Ramos P, Brás JP, Dias C, Bessa-Gonçalves M, Botelho F, Silva J, Silva C, Pacheco-Figueiredo L. Uromonitor: Clinical Validation and Performance Assessment of a Urinary Biomarker Within the Surveillance of Patients With Nonmuscle-Invasive Bladder Cancer. J Urol. 2025 Mar;213(3):304–312. doi: 10.1097/JU.0000000000004335. Epub 2024 Nov 19. PMID: 39561374.

14. Wolff I, Kravchuk AP, Wirtz RM, Schlomm T, Rabien A, Rong D, Hofbauer SL, Labonté FK, Barski D, Otto T, Gössl A, Brookman-May SD, Gilfrich CP, Ecke TH, May M. Real-world performance of Uromonitor® in urothelial bladder cancer detection: a multicentric trial. BJU Int. 2024 Dec;134(6):992–1000. doi: 10.1111/bju.16450. Epub 2024 Jun 24. PMID: 38923777.

15. Ecke TH, Meisl CJ, Schlomm T, Rabien A, Labonté F, Rong D, Hofbauer S, Friedersdorff F, Sommerfeldt L, Gagel N, Gössl A, Barski D, Otto T, Grunewald CM, Niegisch G, Hennig MJP, Kramer MW, Koch S, Roggisch J, Weiß S, Waldner M, Graff J, Veltrup E, Linden F, Hake R, Eidt S, Wirtz RM, Klatte T. Performance of Urinary Markers in Patients With Suspicious Cystoscopy During Follow-up of Recurrent Non-muscle Invasive Bladder Cancer: BTA Stat, NMP22 BladderChek, UBC Rapid Test, CancerCheck UBC Rapid VISUAL, and Uromonitor in Comparison to Cytology. Urology. 2025 Mar;197:119–125. doi: 10.1016/j.urology.2024.11.056. Epub 2024 Dec 1. PMID: 39626834.

16. Rabien A, Rong D, Rabenhorst S, Schlomm T, Labonté F, Hofbauer S, Forey N, Le Calvez-Kelm F, Ecke TH. Diagnostic performance of Uromonitor and TERTpm ddPCR urine tests for the non-invasive detection of bladder cancer. Sci Rep. 2024 Dec 23;14(1):30617. doi: 10.1038/s41598-024-83976-2. PMID: 39715826; PMCID: PMC11666540.

17. OpenAI. GPT-5.2 model documentation [Internet]. San Francisco (CA): OpenAI; 2025 [cited 2025 Dec 24]. Available from: https://platform.openai.com/docs/models

18. Anthropic. Claude model documentation and version registry [Internet]. San Francisco (CA): Anthropic; 2025 [cited 2025 Dec 24]. Available from: https://docs.anthropic.com

19. Google DeepMind. Gemini 3 Pro model card [Internet]. London (UK): Google DeepMind; 2025 [cited 2025 Dec 24]. Available from: https://deepmind.google/technologies/gemini

20. DeepSeek. DeepSeek model documentation and technical reports [Internet]. Beijing (CN): DeepSeek; 2024–2025 [cited 2025 Dec 24]. Available from: https://www.deepseek.com

